# Medical student attitudes to patient involvement in healthcare decision-making and research

**DOI:** 10.1101/2023.03.07.23286892

**Authors:** J. O’Neill, B. Docherty Stewart, A. XN. Ng, Y. Roy, L. Yousif, K. R. McIntyre

## Abstract

**Objective:** Patient involvement is used to describe the inclusion of patients as active participants in healthcare decision-making and research. This study aimed to investigate incoming Year 1 medical (MBChB) students’ attitudes and opinions regarding patient involvement in this context.

**Methods:** We established a staff-student partnership to formulate the design of an online research survey, which included Likert scale questions and three short vignette scenarios designed to probe student attitudes towards patient involvement linked to existing legal precedent. Incoming Year 1 medical students (n = 333) were invited to participate in the survey before formal teaching commenced.

**Results:** Survey data (49 participants) indicate that students were broadly familiar with, and supportive of, patient involvement in medical treatment. There was least support for patient involvement in conducting (22.4%), contributing to (34.7%) or communicating research (30.6%), whereas there was unanimous support for patients choosing treatment from a selection of options (100%).

**Conclusion:** Incoming members of the medical profession demonstrate awareness of the need to actively involve patients in healthcare decision-making but are unfamiliar with the utility and value of such involvement in research. Further empirical studies are required to examine attitudes to patient involvement in healthcare.

Manuscript Revision

This pre-print represents the third version of this manuscript, which has been submitted for peer-review and publication. Further changes are likely to be made before it is published.

**Summary of minor changes made to version 2:**

- Changed of title to focus on ‘healthcare’ research and decision-making.
- Revised the introduction to focus on patient and public involvement (PPI) over ethical themes.
- Changed order of Table 1 and 2 to align with appearance in the text.
- Revised the discussion to focus on PPI, removed of specific results/ percentages, removed references to micro/macro healthcare.
- Reduced and consolidated the strengths and limitations section.
- Edits to general formatting to align with publisher guidelines.
- Added/removed references in line with aforementioned changes to the body of the manuscript.
- Amended spelling (from US English to UK English) and changed to Vancouver Referencing Style in line with publisher guidelines.

## Introduction

Patient and public involvement (PPI) describes active collaboration between patients and/or the wider ‘public’ and researchers. In its broadest sense, PPI gives growing recognition of the need for *inclusive* models of healthcare which view patients as active participants in, rather than passive recipients of, healthcare decision-making and research [1][2].

The historical roots of PPI in healthcare can be traced back to the civil rights movement of the mid-1950s which challenged authoritarianism in favour of democracy [3]. In seeking such democratic input to healthcare, patient support and advocacy groups were formed to challenge traditional forms of medical authoritarianism so that patient and public voices were heard. According to Wilson and colleagues, such was their moral rights as tax-paying ‘consumers’ of the National Health Service (NHS) [3]. In the early 2000s, a statutory ‘Duty to Involve’ patients in policymaking was introduced in England [4] whilst the Health and Social Care Act 2008 required researchers to demonstrate PPI in their work [5][6]. PPI in research may help influence priority setting, experimental design, and future research applications, all of which can improve overall research design [7][8][9]. PPI promotes the patient as an ‘expert in experience’ with their own, unique epistemic value that can enhance research [10]. Patients and the public may be involved in the conduct of, contribution to, and communication of, research. Accordingly, greater patient involvement in healthcare research has been shown to improve health outcomes, mitigate against patient harm and improve patient experiences [11][12].

However, in terms of patient involvement in healthcare decision-making, the move away from beneficent authoritarianism progressed at a far slower pace [13]. It was not until the Supreme Court ruling of *Montgomery v Lanarkshire Health Board* [2015] that greater patient involvement in decisions about their own care was mandated. In her ruling, Lady Hale clarified that patients are no longer to be viewed as “…*passive recipients of the care of the medical profession…*” [14 at s.75]. As a result, there have been noticeable recent moves to adopt models of ‘shared’ and ‘supported’ healthcare decision making [15][16].

To contribute to the existing scholarship on PPI in healthcare decision-making and research, we sought to develop greater appreciation of the pre-conceived attitudes held by incoming members of the medical profession (Year 1 medical students). Our aim was to determine what pre-conceived attitudes these students held towards patient involvement in healthcare.

## Methods

## Design

To ensure effective communication with the study target group (Year 1 medical students), we established a staff-student partnership with four Year 2 medical students from the design phase of the study. The student partners (co-authors BDS, AN, YR, LY) had each recently completed a Student Selected Component (SSC) project on ‘*Patient Perspectives in Research’* supervised by author KM. SSCs are short modules, chosen by students, which allow them to study an area of interest in more depth – a requirement of the General Medical Council (GMC) [17 at s.94]. In completing their SSC, the student partners each conducted a literature review and designed their own PPI project. We adopted a staff-student partnership model reflective of the ‘*student as apprentice’* dynamic described in Olsen’s ‘Student Partnership Framework’ to facilitate the shared pursuit of knowledge and to support development of our student partners’ research skills [18][19].

All authors met on 9^th^ February 2022 to discuss the scope of the project and to outline expected commitments. To establish the study aims and research design, all authors met again on 21^st^ February 2022. During this meeting, staff partners (JO and KM) discussed the background to the study, proposed study aims, and offered potential research questions for student partners to evaluate. The student partners were given time to discuss the proposed project amongst themselves and were provided whiteboard pens, post-it notes and paper to record their thoughts and amendments.

At the end of these discussions, authors agreed to conduct an exploratory research survey with the research aim of examining incoming medical students’ attitudes to patient involvement in healthcare decision-making and research. Student partners were interested in exploring student perceptions at all levels of academic study, however pragmatic constraints (e.g. academic research time), led us to focus on incoming medical students for this initial study. A further outcome of this meeting was the suggestion to contextualise some statements in the research survey (for example *“doctors should work in partnership with patients”*) with a scenario. Subsequently, JO developed three scenarios to directly reflect current legal precedent and relevant regulatory issues pertaining to patient involvement in healthcare (Table 1). Our study took place before the Supreme Court ruling in *McCulloch v Forth Valley Health Board* [2023] which clarified that determination of ‘reasonable treatment alternatives’ is a matter of professional judgment which does not need to include the patient [20]. Accordingly, the earlier ruling in *Montgomery* which held that patients be informed of the benefits, material risks and reasonable treatment options was used in our scenarios [14]. Following the initial draft, KM and student partners advised on and edited the wording of the scenarios. An iterative feedback loop was established thereafter, whereby all authors discussed and edited the research survey until a consensus was reached.

**Table 1.** Three ethical scenarios were drafted in collaboration with student partners and were designed to interrogate the students’ views related to existing legal precedent (‘rationale’). The scenarios and corresponding options are provided in the table, as they appeared in the survey, alongside participants’ responses (n = 47).

| Scenario | Rationale | Scenario and survey response options | Response<br>[No. (% total)] |
| --- | --- | --- | --- |
| 1 | Representative of the case of *Montgomery v Lanarkshire* [2014] - which holds that patients must be informed of the benefits, material risks and reasonable treatment alternatives [at 87; 90] – scenario 1 describes the case of an expectant mother with type 1 diabetes mellitus due to deliver a larger than average baby. Participants were asked whether doctors should inform the mother of risks and alternative treatment options. | An expectant mother known to have type 1 diabetes mellitus is admitted to the labour ward and is expected to give birth soon. The mother, who is also of short stature, asks doctors whether there are any risks that she should be made aware of. Doctors know that since the mother has diabetes mellitus, she is more likely to be carrying a larger than average baby and that there may be several known complications associated with giving birth naturally compared to via a caesarean section. What should the doctors do? |  |
|  |  | Do not tell the patient of the risks and make a decision on her behalf | 0 (0.0) |
|  |  | Tell the patient of the risks and any options available | 47 (100.0) |
| 2 | Representative of the well-established Torts of Negligence and Battery as they relate to medical consent (see, for example, *Schoendorff v Society of New York Hospital* 1914; *Bolam v Friern Hospital Management Committee*, 1957). Participants were asked whether a surgeon undertaking surgery on a patient for a specified purpose could then undertake an additional, non-urgent procedure without explicit consent. | A patient consents to undergo minor surgery to have a mole removed. During the surgical procedure, however, doctors discover that the patient requires an additional (non-urgent) surgical intervention. This was not previously discussed with the patient. What should the doctors do? |  |
|  |  | Proceed with the additional surgical intervention | 3 (6.4) |
|  |  | Waken the patient and ask for their consent before continuing | 44 (93.6) |
| 3 | Representative of the recommendations made in *First Do No Harm* relating to increased patient input to innovative medicines and medical devices development (Cumberlege, 2020, see ss.2.135, 2.81, 2.107). Participants were asked whether researchers should take patients' lived experience into account or rely solely upon scientific clinical trial evidence during evaluation. | A group of doctors are involved in pioneering a new surgical technique using a new implant. Some months after the initial surgical implantations were performed, patients begin to complain of painful side effects. This is in spite of the fact that clinical trial data is strongly in favour of this new technique and shows little, to no, side-effects. What should the doctors do with this information from their patients? |  |
|  |  | Disregard this information and continue to rely upon the scientific clinical trial evidence to provide the implantation for new patients | 0 (0.0) |
|  |  | Explore the existing patients' experiences in more depth and use this to shape future practice | 47 (100.0) |

## Ethical approval

Approval for this study was obtained from the College of Medical, Veterinary and Life Sciences Ethics Committee for Non-clinical Research Involving Human Participants [No: 200210131] with staff and student partners included as named researchers. The ensuing collection, storage and processing of personal data was in accordance with the Data Protection Act 2018 [21].

## Recruitment

The survey (Supplementary File 1) was built and disseminated via Qualtrics (Qualtrics LLC 2022) using an institutional license. Incoming Year 1 medical students (n = 333) were invited to participate in the research study on 2^nd^ September 2022 via an announcement posted on the institutional virtual learning environment, Moodle. An additional reminder was posted on 21^st^ September 2022. The release of the survey in this manner, prior to the start of the academic year (20^th^ September 2022), was made possible on account of our well-established mandatory online pre-entry induction course which all Year 1 medical students gain access to before formal teaching begins [22][23]. A pragmatic decision was taken to close the survey on 22^nd^ September 2022 before students’ first scheduled lecture on the topic of ethics to remove the potential for influence of this teaching on the study results. In this small-scale study, our aim was to gather an insight into the attitudes of new members of the medical profession at our institution, rather than to generalise to a broad population.

## Results

Responses were disregarded from analyses where only demographic information was given (n = 6). Subsequently, three partially complete and 46 complete responses were included in the analyses (49 total, 14.7% of year group). Most participants were female (40/49, 81.6%), between 18-21 years old (36/49, 73.5%) and had joined the University directly from school (36/49, 73.5%). Demographic characteristics of participants are summarised in Table 2.

**Table 2.** Demographic characteristics of survey participants.

| Characteristic | No. (% total) |
| --- | --- |
| **Gender** |  |
| Female | 40 (81.6) |
| Male | 9 (18.4) |
| Not listed | 0 (0.0) |
| Prefer not to say | 0 (0.0) |
| **Age** |  |
| 17 or under | 9 (18.4) |
| 18-21 | 36 (73.5) |
| 22-25 | 3 (6.1) |
| 26-29 | 1 (2.0) |
| 30+ | 0 (0.0) |
| Prefer not to say | 0 (0.0) |
| **Educational background** |  |
| Foundation course or pre-med | 3 (6.1) |
| Gap year | 4 (8.2) |
| Graduate | 4 (8.2) |
| Repeating or returning to Year 1 | 2 (4.1) |
| School leaver | 36 (73.5) |
| Other | 0 (0.0) |

Survey participants were asked to rate how much they agreed or disagreed with seven statements on a five-point Likert scale (Figure 1). Most students (35/49, 71.4%) agreed or strongly agreed that patients’ lived experience should be valued as much as clinical knowledge or expertise. All participants agreed that it is important to represent the patient voice in healthcare (4/49, 8.2% agreed; 45/49, 91.8% strongly agreed), to consider patient input alongside scientific findings (23/49, 46.9% agreed; 26/49, 53.1% strongly agreed) and to involve patients in management decisions (9/49, 18.4% agreed; 40/49, 81.6% strongly agreed). Conversely, four participants (8.2%) agreed that a doctor should have the final say in decision-making and some participants said that patients should be the passive recipients of care (3/49, 6.1% agreed; 1/49, 2.0% strongly agreed).

**Figure 1.**
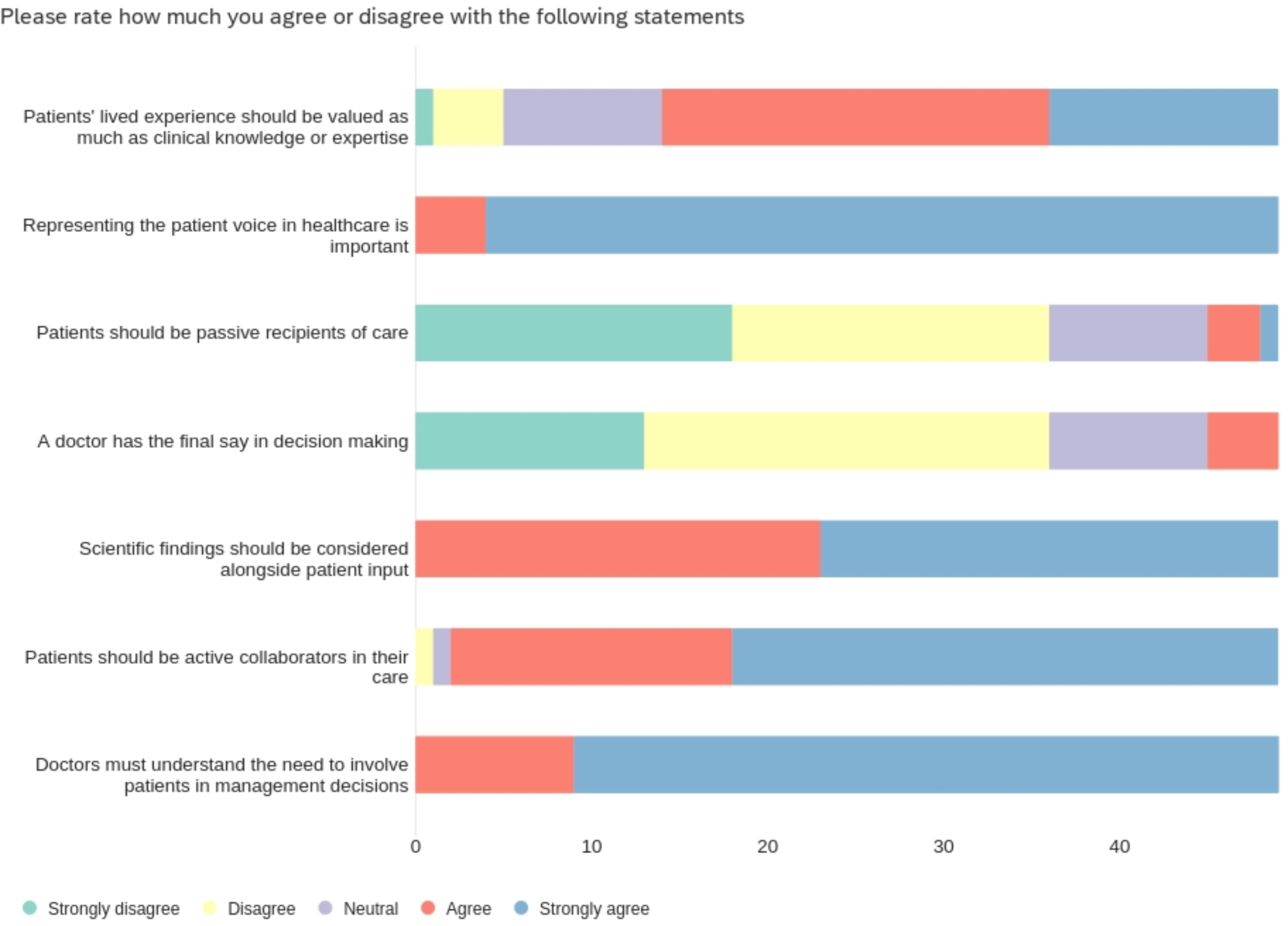
Participants’ responses to Likert scale statements (n = 49).

Participants were presented with three scenarios related to an existing legal precedent of which the participants were not informed and were asked to choose what the doctors should do in each scenario. All participants (n = 47) chose the option that aligned with the legal precedent for scenarios 1 and 3 (Table1). In scenario 2, participants were asked whether a surgeon undertaking surgery on a patient for a specified purpose could then undertake an additional, non-urgent procedure without explicit consent. Three participants (6.4%) opted to proceed with the additional surgical intervention in this scenario (Table 1).

Participants were asked to select aspects of healthcare that they thought patients could be involved in from a standard list (Table 3). They could select multiple options. The most popular options were ‘choosing treatment from a selection of options’ (46/46, 100% of participants) and ‘deciding where to receive healthcare’ (41/46, 89.1%). Fewer participants selected options related to designing (17/46, 37.0%), conducting (11/46, 23.9%) or communicating medical research findings (15/46, 32.6%).

**Table 3 1.** Ways for patients to be involved in healthcare. Survey participants (n = 46) could select multiple options.

| Which aspects of healthcare do you think patients can be involved in? | No. of responses (% total) |
| --- | --- |
| Choosing treatment from a selection of options | 46 (100) |
| Communicating research findings | 15 (32.6) |
| Conducting research | 11 (23.9) |
| Contributing to the design of research studies | 17 (37.0) |
| Deciding where to receive healthcare | 41 (89.1) |
| Medical decision-making about treatment | 30 (65.2) |
| None of the above | 0 (0.0) |

To explore participants’ own experiences, participants were asked whether they felt their opinion had been valued in their own experience of healthcare. Closed text responses (Table 4) indicated that most participants felt that their opinion had been valued (29/38, 63.0%). Nine participants (19.6%), all female, felt that their opinion had not been valued.

**Table 4 2.** Participants were asked to disclose whether they felt their opinion has been valued in their own experience of healthcare (n = 46).

| In your personal experience with healthcare, have you felt that your opinion has been valued? | No. female (% female) | No. male (% male) | No. overall respondents (% total) |
| --- | --- | --- | --- |
| Yes | 23 (60.5) | 6 (75.0) | 29 (63.0) |
| No | 9 (23.7) | 0 (0.0) | 9 (19.6) |
| Not applicable | 6 (15.8) | 2 (25.0) | 8 (17.4) |

## Discussion

To the best of our knowledge, this is the first study in the UK to examine medical student attitudes towards patient involvement in healthcare decision-making and research. Our findings demonstrate that participants were broadly supportive of patient involvement in key aspects of medical decision-making yet less supportive of research involvement.

## Patient Involvement in Research

The NIHR defines patient involvement in research as “*research being carried out ‘with’ or ‘by’ members of the public rather than ‘to’, ‘about’ or ‘for’ them*” [24]. This reflects a model of shared decision-making whereby the epistemic value of the patient is recognised at the research and development stage of product development [2]. Our data indicate less support, in a new medical student cohort, regarding general patient involvement in research compared to healthcare decision-making. Students were less aware of opportunities to involve patients in conducting, contributing to, and communicating research. Our findings are reflective of a systematic review by Biddle et al., (2020) which found growing support for the concept of patient involvement in healthcare but “*to a lesser extent, in health research*” [25 at p24]. Biddle and colleagues describe the UK as a leading contributor to patient involvement in research compared to several other European countries [25]. They note that, whilst the *“general attitude towards … [patient involvement…] is changing”* to reflect more acceptance, it is often in a *“marginal or tokenistic”* sense [25 at p24]. Similarly, a study by Boaz et al., (2016) suggests that researchers demonstrate *“active resistance”* [26 at p600] to sharing control of the research process with patients constrained to “*tinker[ing] at the edges*” [26 at p592]. Such attitudes appear to be reflected in our incoming medical student cohort, the majority of which did not agree that patients can be involved in research processes (Table 3). It is vital to address why attitudes towards patient involvement in the research process is less agreeable. These combined findings indicate that there may be a need to improve student awareness of the role, and benefits, of patient involvement in research.

## Patient Involvement in Healthcare Decision-Making

In healthcare, shared decision-making is a complex process in which doctors diagnose the patient and then select appropriate treatment options for presentation to the patient. The initial selection of treatment is viewed as a matter of professional judgement which does not involve the patient [13][20]. Over a third of research participants did not indicate support for patient involvement in ‘medical decision-making about treatment’ (Table 3). For the purposes of informed consent according to *Montgomery*, patients should be informed of the reasonable treatment options [14]. Our participants demonstrated strong support for such patient choice from a range of treatment options. The General Medical Council (GMC) recognise that “*[d]ecision making is an ongoing process focused on meaningful dialogue*” [15 at p7]. Accordingly, once a treatment plan has been decided upon, the patient should continue to be informed as the condition is managed. There was unanimous agreement amongst our participants that doctors *should* involve patients in such treatment ‘management decisions’ (Figure 1). Further support for this premise may be derived from our participants’ strong support for the patient voice in healthcare which demonstrates respect for *inclusivity* and active participation in healthcare decision-making.

Overall, our incoming medical students demonstrated supportive attitudes towards patient involvement in informed, medical decision-making with most *against* doctors having the final say in decision-making and of patients being viewed as passive recipients of care. Such pre-conceived attitudes may derive from a clear sense of patients as consumers, and the growing role of consumerism in healthcare [27]. Responses to the scenarios (Table 1), which reflected key legal principles, may offer supportive evidence of this premise. Participants held unanimously supportive attitudes towards basic principles of consent, as per *Montgomery v Lanarkshire Health Board* [2015][14]. A small proportion of participants, however, demonstrated support for the performance of surgical procedures beyond the scope of initial consent, which fails to adequality involve and inform the patient. It is, therefore, important that incoming members of the profession are made aware of the dynamic and *ongoing* nature of consent during medical training, such as through vocational skills or clinical placement teaching.

## Strengths and Limitations

We acknowledge that social desirability bias (SDB) – defined by Zerbe and Paulhus (1987) as *“the tendency of individuals to present themselves favourably with respect to current social norms and standards”* – may have led our participants to respond in a manner ‘expected of them’ [28 at p250]. However, we recognise that as incoming medical students, our participants would have been expected to demonstrate basic awareness of pertinent medical issues in their medical applications and interviews and are therefore likely to have had pre-existing knowledge that influenced their attitudes to PPI in a positive manner. Additionally, high non-response rates in voluntary surveys are well-documented. Indeed, Porter and Whitcomb (2005) suggest that the decision *not* to participate in a survey is multi-factorial; developing an appreciation of these factors may assist in interpreting the *quality* of data and could be explored in future work [29].

A key strength of our study lies in the involvement of medical students as active partners in our research team, which ensured appropriate communication with our target participants. *Strength* also derives from our inclusion of questions on demographics which enable us to draw hypotheses on the specific factors associated with response decisions. Most participants were ‘school leavers’ under the age of 21, which is broadly reflective of the cohort demographic [30]. The notably higher response rate from female medical students is not surprising given that the GMC reported that most medical entrants were women in 2022 [30]. Female medics are also more likely to spend time engaging in meaningful discussions with patients [31][32]. Such gender disparity may be linked to female egalitarianism or, indeed, the lived experiences of female medics as patients themselves, given the significantly negative correlations between female gender and integration into shared decision-making [33][34]. Indeed, fewer of our female participants felt that *their* opinion was been valued in their personal healthcare experiences compared to males. Whilst we may we conclude that interest in patient involvement positively correlates with survey engagement, we may *conversely* assume that some non-participants may have found the subject matter to be ‘*boring*’ or to lack relevance which may have utility for medical curriculum development to ensure students are engaged on the subject [29 at p129].

## Future Research

The evolution of student attitudes towards PPI is something that we are collectively keen to explore further. There is evidence to suggest that medical student attitudes can be shaped by their experiences in the clinical environment and from exposure to the negative attitudes of practicing clinicians [35]. Such research may align with existing work which suggest that empathy – *“one of the most highly desirable professional traits”* and *“crucial for [establishing the] successful physician-patient”* relationship that underpins meaningful patient involvement – erodes over time [36 at p244].

## Conclusion

Incoming medical students demonstrate awareness of the need for patient involvement in healthcare treatment but lack appreciation for the role of patient involvement in medical research, despite the long-established history of PPI in research. Further empirical studies are required to determine whether such favourable attitudes to patient involvement wane over time. We anticipate that our collective findings may serve as the basis for future research and may have utility for promoting ongoing medical education to promote the value of patient involvement in medicine and health research.

## Data Availability

All data produced in the present work are available upon reasonable request to the authors.

## Supplementary File 1: Survey

### Student perceptions of patients’ involvement in medical care

**Start of Block: Participant Information**

Q1 **Participant Information**

You are being invited to take part in a research study. Before you decide, it is important for you to understand why the research is being done and what it will involve. Please take time to read the following information carefully and discuss it with others if you wish. Ask us if there is anything that is not clear or if you would like more information. By completing the questionnaire you will be considered to be consenting to the study.

**Background to this study**

There is growing interest in the role and scope of patient involvement in healthcare in the United Kingdom, however, research is still ongoing to reach a consensus as to what this means in practice.

The purpose of this study is to investigate medical students’ attitudes and perceptions of patient involvement in healthcare. It aims to do so by surveying incoming year 1 medical students. This study has been designed in collaboration with four current year 3 medical students.

**Why am I being asked to participate?**

You have been asked to take part as you are a current 1st year medical student. It is your decision whether or not to take part in this study. A decision not to participate will not affect your grades in any way.

**What will happen if I decide to take part?**

If you take part, you will be asked to fill out a short online questionnaire. The questionnaire will take no longer than 15 minutes to complete. The completed questionnaires will be analyzed to see if there are any common themes. The information we gather will give us a better understanding of how students view patient involvement in healthcare and may help in future course design.

**Are there any benefits or risks involved?**

Although there is no specific benefit to taking part in the study, completing the questionnaire may allow you to reflect on your experiences, which you may find helpful. In the unlikely event that participants inadvertently disclose personal information such as that regarding the mental and/or physical health of themselves or others, information relating to criminal acts and/or acts of professional misconduct, participants should understand that the research team may have to report such disclosures to the appropriate authorities as deemed necessary by the nature of the disclosure.

**What will happen to my data if I take part?**

Researchers from the University of Glasgow collect, store and process all personal information in accordance with the General Data Protection Regulation (2018). You will not be asked to disclose any personally identifiable information. All data will be stored in electronic format on secure password-protected computers. The data will be stored in archiving facilities in line with the University of Glasgow retention policy of up to 10 years. After this period, further retention may be agreed or your data will be securely destroyed in accordance with the relevant standard procedures. Your rights to access, change or move the information we store may be limited, as we need to manage your information in specific ways in order for the research to be reliable and accurate. You can find out more about how we use your information from Dr Jennifer O’Neill.

**How will the results be communicated?**

It is anticipated that the results of the study will be presented both internally and externally and submitted for publication in the appropriate literature. No-one will be identifiable from the information presented. The project has been reviewed by the College of Medicine, Veterinary and Life Sciences Ethics Committee.

If you have any questions or concerns about the research, you can contact the organizer of the study: Dr Jennifer O’Neill, (Lecturer in Biology) by

Thank you for taking time to read this information sheet.

Q3 I understand that should I inadvertently disclose personal information such as that regarding the mental and/or physical health of myself or others, information relating to criminal acts and/or acts of professional misconduct, the research team may have to report such disclosures to the appropriate authorities as deemed necessary by the nature of the disclosure.

Yes (1) No (2)

*Skip To: End of Survey If I understand that should I inadvertently disclose personal information such as that regarding the… = No*

Q4 I have read the participant information sheet and understand that by completing the questionnaire I consent to participation.

Yes (1) No (2)

*Skip To: End of Survey If I have read the participant information sheet and understand that by completing the questionnaire… = No*

**End of Block: Participant Information**

**Start of Block: Demographics**

Q5 What is your gender?

Female (1) Male (2) Not listed (3) Prefer not to say (4)

Q6 What age are you?

17 or under (1) 18-21 (2) 22-25 (3) 26-29 (4) 30+ (5) Prefer not to say (6)

Q7 What is your educational background

Foundation course or pre-mend (1) Gap year (2) Graduate (3) Repeating or returning to year 1 (4) School leaver (5) Other (6)

**End of Block: Demographics**

**Start of Block: Patient involvement**

Q8 **This section of the survey aims to explore your views on the roles of doctors and patients’ in healthcare.**

Q9 Please rate how much you agree or disagree with the following statements

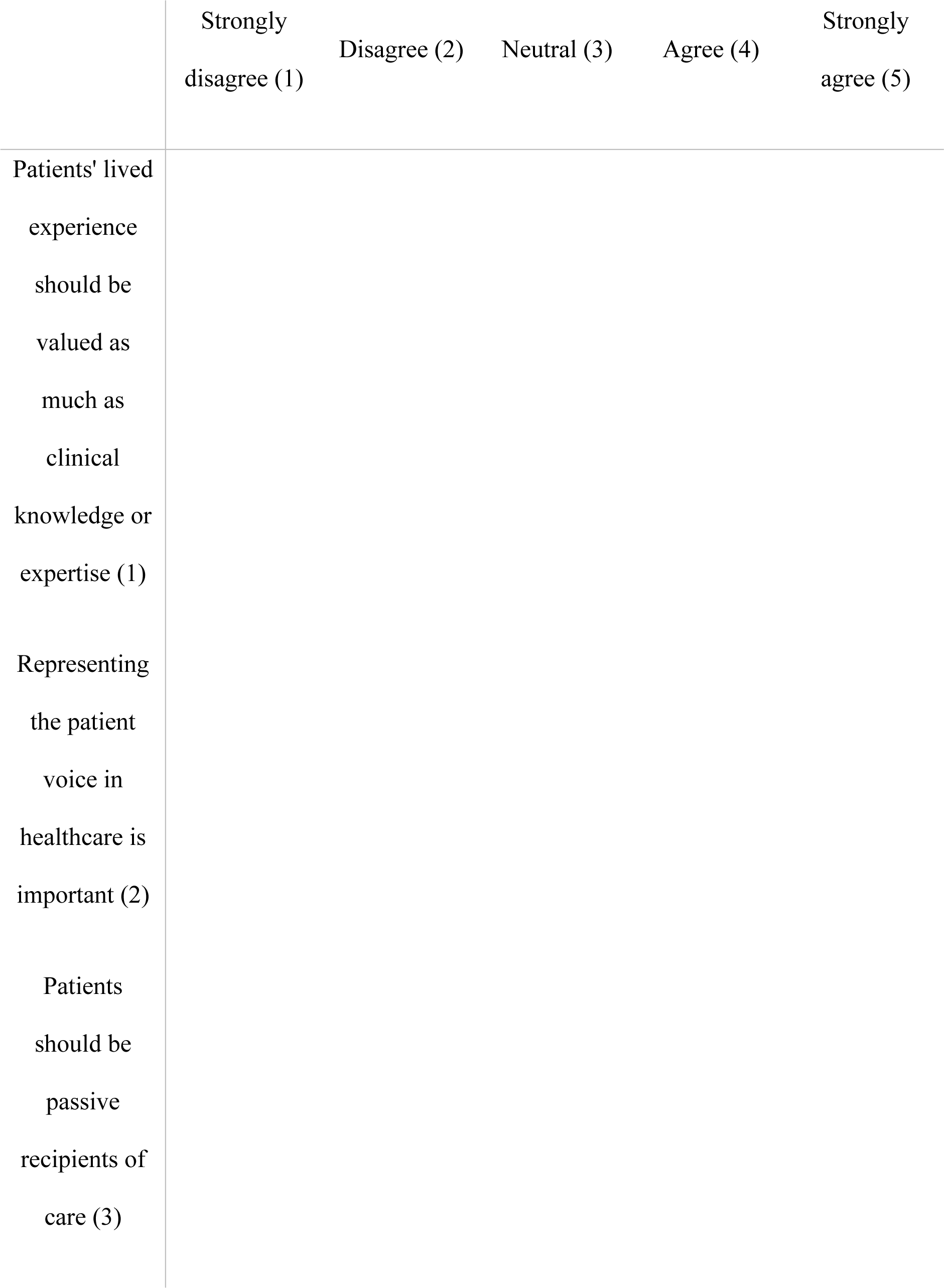
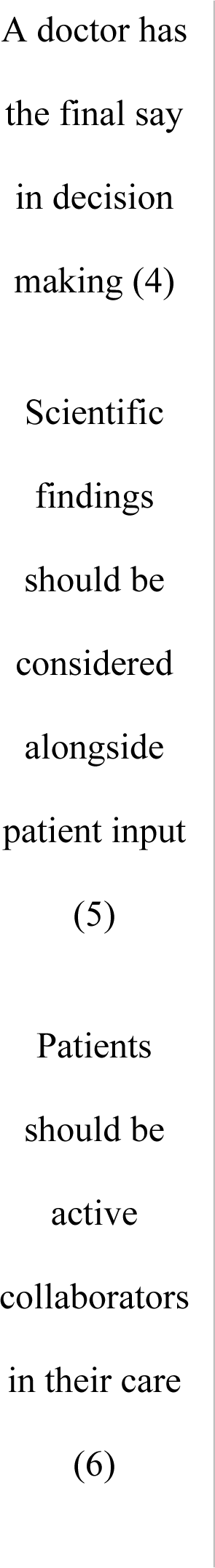
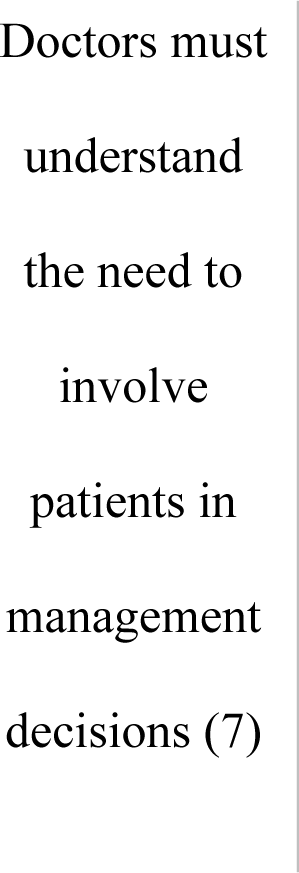

**End of Block: Patient involvement**

**Start of Block: Patient scenarios**

Q10 **Patient scenarios**

For each scenario, select the option that you think best suits the situation.

Q11 **Scenario 1**

Q12 An expectant mother, known to have type 1 diabetes mellitus, is admitted to the labor ward and is expected to give birth soon. The mother, who is also of short stature, asks doctors whether there are any risks that she should be made aware of. Doctors know that since the mother has diabetes mellitus, she is more likely to be carrying a larger than average baby and that there may be several known complications associated with giving birth naturally compared to via a caesarean section. What should the doctors do?

Do not tell the patient of the risks and make a decision on her behalf (1) Tell the patient of the risks and any options available (2)

Q14 **Scenario 2**

Q13 A patient consents to undergo minor surgery to have a mole removed. During the surgical procedure, however, doctors discover that the patient requires an additional (non-urgent) surgical intervention. This was not previously discussed with the patient. What should the doctors do?

Proceed with the additional surgical intervention (1) Waken the patient and ask for their consent before continuing (2)

Q15 **Scenario 3**

Q16 A group of doctors are involved in pioneering a new surgical technique using a new implant. Some months after the initial surgical implantations were performed, patients begin to complain of painful side effects. This is in spite of the fact that clinical trial data is strongly in favour of this new technique and shows little, to no, side-effects. What should the doctors do with this information from their patients?

Disregard this information and continue to rely upon the scientific clinical trial evidence to provide the implantation surgery for new patients (1) Explore the existing patients’ experiences in more depth and use this to shape future practice (2)

**End of Block: Patient scenarios**

**Start of Block: Patient involvement**

Q17 Which aspects of healthcare do you think patients can be involved in? (select all that apply)

▢ Choosing treatment from a selection of options (1) ▢ Communicating research findings (2) ▢ Conducting research (3) ▢ Contributing to the design of research studies (4) ▢ Deciding where to receive healthcare (5) ▢ Medical decision-making about treatment (6) ▢ None of the above (7)

Q18 In your personal experience with healthcare, have you felt that your opinion has been valued?

Yes (1) No (2) Not applicable (3)

**End of Block: Patient involvement**

**Start of Block: Free text questions**

Q19 Please use this space to provide any other relevant comments. We do not wish to capture any personally identifiable information or information related to personal circumstances. Students are reminded to avoid sharing such information when responding to this question.

________________________________________________ ________________________________________________ ________________________________________________ ________________________________________________ ________________________________________________

**End of Block: Free text questions**

## References

[1] National Institute for Health Research (NIHR) School for Primary Care Research (2022) ‘What is patient and public involvement and public engagement?’ Accessed at https://www.spcr.nihr.ac.uk/PPI/what-is-patient-and-public-involvement-and-engagement on 19 July 2023

[2] O’Neill J. Towards conjoint solidarity, Bioethics 2021; 36(5): 535-546. https://doi.org/10.1111/bioe.12940

[3] Wilson P, Mathie E, Keenan J, McNeilly E, Goodman C, Howe, A, Poland F, Staniszewska S, Kendall S, Munday D, Cowe M, Peckham S. ‘ReseArch with Patient and Public invOlvement: a RealisT evaluation – the RAPPORT study’. Southampton (UK): NIHR Journals Library; 2015 https://doi.org/10.3310/hsdr03380

[4] National Health Service Act 2006, s.252 as amended by the Local Government & Public Involvement in Health Act 2007

[5] Health and Social Care Act 2008

[6] Edwards V, Wyatt K, Logan S, Britten N. Consulting parents about the design of a randomized controlled trial of osteopathy for children with cerebral palsy. Health Expect. 2011; 14: 429–38. https://doi.org/10.1111/j.1369-7625.2010.00652.x

[7] Rolfe DE, Ramsden VR, Banner D, Graham ID. Using qualitative Health Research methods to improve patient and public involvement and engagement in research. Res Involv. 2018; 4(49) https://doi.org/10.1186/s40900-018-0129-8

[8] Andrews LM, Allen H, Sheppard ZA, Baylis G, Wainwright TW. More than just ticking a box…how patient and public involvement improved the research design and funding application for a project to evaluate a cycling intervention for hip osteoarthritis. Res Involv. 2015; 1(13) https://doi.org/10.1186/s40900-015-0013-8

[9] Sacristán JA, Aguarón A, Avendaño-Solá C, Garrido P, Carrión J, Gutiérrez A, Kroes R, Flores A. Patient involvement in clinical research: why, when, and how. Patient Prefer Adher. 2016; 27(10): 631–40 https://doi.org/10.2147/PPA.S104259

[10] Howlett H, Catterick M, Charlotte **, Karen **, Cheyenne **, Onukwugha F, Roberts H, Dyson J, Smith L. Reflections of experts by experience and research team members on research and development about a sensitive issue that attracts stigma. Research for All. 2023; 7(1): 2 https://doi.org/10.14324/RFA.07.1.02

[11] Bombard Y, Baker GR, Orlando E, Fancott C, Bhatia P, Casalino S, Onate K, Denis J, Pomey M P. Engaging patients to improve quality of care: a systematic review. Implemenat.Sci. 2018; 13(98). https://doi.org/10.1186/s13012-018-0784-z

[12] World Health Organization. ‘WHO calls for urgent action to reduce patient harm in healthcare’. 13 Sept. 2019. Accessed at https://www.who.int/news/item/13-09-2019-who-calls-for-urgent-action-to-reduce-patient-harm-in-healthcare on 19 July 2023

[13] Bolam v Friern Hospital Management Committee [1957] 1 WLR 583 as per Justice McNair at 586

[14] Montgomery v Lanarkshire Health Board [2015] UKSC 11

[15] General Medical Council (2020) ‘Guidance on professional standards and ethics for doctors: Decision making and consent. The Seven Principles of Decision Making and Consent: Two’. 9 Nov. 2020. Accessed at https://www.gmc-uk.org/-/media/documents/updated-decision-making-and-consent-guidance_pdf-84160128.pdf on 19 July 2023

[16] Royal College of Surgeons of England (2018) ‘Consent: Supported Decision Making’. Nov 2018. Accessed at https://www.rcseng.ac.uk/standards-and-research/standards-and-guidance/good-practice-guides/consent/ on 19 July 2023

[17] General Medical Council. ‘Tomorrow’s Doctors: outcomes and standards for undergraduate medical education’. Sept. 2009. s.94, p.50. Accessed at http://www.ub.edu/medicina_unitateducaciomedica/documentos/TomorrowsDoctors_2009.pdf on 19 July 2023

[18] Holen R, Ashwin P, Maassen P, Stensaker B. Student partnership: exploring the dynamics in and between different conceptualizations. Stud.High.Educ. 2020; 46(12): 2726–2737 https://doi.org/10.1080/03075079.2020.1770717

[19] Olsen J P. The institutional dynamics of the European university. In: University Dynamics and European Integration, edited by P. Maassen and J.P. Olsen; 2007. pp25-54. Dordrecht: Springer. https://doi.org/10.1007/978-1-4020-5971-1_2

[20] McCulloch v Forth Valley Health Board [2023] UKSC 26

[21] Data Protection Act (U.K) 2018

[22] McIntyre K. Bridging the gap: implementation of an online induction course to support students’ transition into first year medicine. MedEdPublish. 2021; 9(1): 193 https://doi.org/10.15694/mep.2020.000193.2

[23] McIntyre K, O’Neill J. The process of adapting an online induction course to support distinct student cohorts. JLDHE. 2022: 24 https://doi.org/10.47408/jldhe.vi24.829

[24] National Institute for Health Research (NIHR) Involve (n.d.) What is public involvement in research? Accessed at https://www.invo.org.uk/find-out-more/what-is-public-involvement-in-research-2/ on 19 July 2023

[25] Biddle MSY, Gibson A, Evans D. Attitudes and approaches to patient and public involvement across Europe: A systematic review. Health Soc. Care Community. 2020; 29(1): 18–27 https://doi.org/10.1111/hsc.13111

[26] Boaz A, Biri D, McKevitt C. Rethinking the relationship between science and society: Has there been a shift in attitudes to patient and public involvement and public engagement in science in the United Kingdom? Health Expect. 2016; 19(3): 592–601 https://doi.org/10.1111/hex.12295

[27] Latimer T, Roscamp J, Papanikitas A. Patient-centredness and consumerism in healthcare: an ideological mess. J R Soc Med. 2017; 110(11): 425–427. https://doi.org/10.1177/0141076817731905

[28] Zerbe WJ, Paulhus DL. Socially desirable responding in organizational behavior: A re-conception. AMR. 1987; 12(2). https://doi.org/10.5465/amr.1987.4307820

[29] Porter SR, Whitcomb ME. Non-response in student surveys: The role of demographics, engagement and personality. Res High Educ. 2005; 46(2) https://doi.org/10.1007/s11162-004-1597-2

[30] General Medical Council. ‘The state of medical education and practice in the UK: The workforce report 2022’. Accessed at https://www.gmc-uk.org/-/media/documents/workforce-report-2022---full-report_pdf-94540077.pdf on 19 July 2023

[31] Alameddine M, Otaki F, Bou-Karroum K, Du Preez L, Loubser P, AlGurg R, Alsheikh-Ali A. Patients’ and physicians’ gender and perspective on shared decision-making: A cross-sectional study from Dubai. PLoS ONE. 2022; 17(9): e0270700 https://doi.org/10.1371/journal.pone.0270700

[32] Roter DL, Hall JA. Why physician gender matters in shaping the physician-patient relationship. J. Women’s Health. 2009; 7(9) https://doi.org/10.1089/jwh.1998.7.1093

[33] Araújo EB, Araújo NAM, Moreira AA, Herrmann HJ, Andrade JS. Gender differences in scientific collaborations: Women are more egalitarian than men. PLoS One. 2017; 12(5): e0176791 https://doi.org/10.1111/hsc.13111

[34] Ommen O, Theum S, Janssen C. The relationship between social support, shared decision-making and patient’s trust in doctors: a cross-sectional severe of 2,197 inpatients using the Cologne Patient Questionnaire. Int. J. Pub Health.2011; 56: 319*–*327

[35] Pohontsch NJ, Stark A, Ehrhardt M, Kotter T, Scherer M. Influences on students’ empathy in medical education: an exploratory interview study with medical students in their third and last year. BMC Med. 2018; 18(231) https://doi.org/10.1186/s12909-018-1335-7

[36] Newton B, Barber L, Cleveland E, O’Sullivan P. Physician-patient relationship: Is there hardening of the heart during medical school? Acad. Med. 2008; 83(3): 244–249 https://doi.org/10.1097/ACM.0b013e3181637837

